# Human genetic investigation of TMPRSS6 as a therapeutic target for liver disease: efficacy and safety liabilities

**DOI:** 10.64898/2026.10.29.26366887

**Authors:** Marie-Joe Dib, Marijana Vujkovic, Stephen Burgess, Tithi Dutta, Ioanna Tzoulaki, Abbas Dehghan, Hanieh Yaghootkar, Devendra Meena

## Abstract

**Background & Aims:** Disease-modifying therapies for steatohepatitis and fibrosis remain limited. Matriptase-2 (TMPRSS6), a serine protease suppressing hepcidin to regulate iron homeostasis, is under clinical investigation for iron overload, and preclinical evidence suggests its inhibition may attenuate metabolic liver disease. Whether these effects translate to humans remains unknown.

**Methods:** We proxied TMPRSS6 inhibition using the missense variant rs855791, weighted on circulating hepcidin. Using Mendelian randomization (MR), we assessed effects on (1) magnetic resonance imaging-derived liver phenotypes, including corrected T1 (cT1) and proton density fat fraction (PDFF), liver enzymes and clinical liver outcomes; (2) cardiometabolic endpoints; (3) 2,940 plasma proteins; (4) clinical biomarkers; and (5) 2,272 disease endpoints. Colocalization and mediation analyses were used to characterize mechanisms.

**Results:** Genetically proxied TMPRSS6 inhibition was associated with lower liver iron content (β = −2.50, P<0.001) and higher cT1 (β = 1.59, P<0.001), with colocalization indicating a shared causal signal. No association was observed with PDFF, liver volume, NAFLD, cirrhosis or hepatocellular carcinoma. Genetically proxied TMPRSS6 inhibition was also associated with lower C-reactive protein (β = −0.17, P<0.001), higher LDL cholesterol (β = 0.22, P<0.001) and higher coronary artery disease (CAD) risk (OR = 1.59, P<0.001), with weak colocalization of CAD with hepcidin (PP.H4 = 0.56) and LDL cholesterol (PP.H4 = 0.48). Mediation analysis suggested LDL cholesterol may account for approximately 21% (95% CI 9% to 34%) of the CAD association.

**Conclusions:** Human genetic evidence supports hepatic iron lowering by TMPRSS6 inhibition, but not an effect on steatosis. Higher cT1, without concordant associations with clinical liver outcomes, may reflect reduced hepatic iron rather than worsening fibroinflammation. The CAD signal, partly mediated by LDL cholesterol, warrants careful consideration in trial design.

**Impact and implications:** Preclinical studies have identified TMPRSS6 inhibition as a potential therapeutic strategy for metabolic liver disease, but its effects in humans and potential safety consequences remain uncertain. Using human genetic evidence, we show that genetically proxied TMPRSS6 inhibition is associated with lower hepatic iron content but not hepatic steatosis, while identifying increased coronary artery disease risk as a potential cardiovascular safety liability. These findings are relevant to researchers and drug developers pursuing TMPRSS6-directed therapies, as they suggest that clinical benefit may be more likely in patients with excess hepatic iron and highlight the importance of cardiovascular risk assessment and lipid monitoring during clinical development. As genetic proxies reflect lifelong perturbation rather than pharmacological treatment, prospective clinical studies are required to determine whether these hepatic and cardiovascular effects are reproduced with therapeutic TMPRSS6 inhibition.

**Highlights:**

- Genetically predicted TMPRSS6 inhibition was associated with lower hepatic iron content.
- No associations were observed with hepatic steatosis or liver volume.
- Genetically predicted TMPRSS6 inhibition was associated with increased CAD risk.
- LDL-C mediated around one fifth of the CAD association.

## Introduction

Liver disease accounts for over two million deaths annually, representing 4% of all deaths worldwide (1). Metabolic dysfunction-associated steatotic liver disease (MASLD) is the most prevalent chronic liver disease globally, affecting an estimated 25-38% of adults with projections anticipating continued increases in metabolic dysfunction-associated steatohepatitis (MASH)-related cirrhosis, hepatocellular carcinoma, and liver-related mortality through 2030 (2). Despite this burden, disease-modifying therapies that reliably halt or reverse steatohepatitis and fibrosis remain limited, highlighting the need to identify and validate novel therapeutic targets.

TMPRSS6 (Transmembrane Protease Serine 6), also known as matriptase-2, is a type II membrane-bound serine protease predominantly expressed in hepatocytes, that plays a critical role in systemic iron homeostasis (3,4). TMPRSS6 cleaves the hemojuvelin co-receptor, thereby attenuating bone morphogenetic protein (BMP) signaling and limiting hepcidin transcription (5). In preclinical MASLD models, hepatocyte-directed downregulation of Tmprss6 using a GalNAc-conjugated antisense approach reduced steatohepatitis and attenuated fibrosis (6). TMPRSS6 inhibition is currently in drug development pipelines for iron overload disorders, with early-phase clinical studies providing initial evidence of tolerability (7). However, whether the hepatic benefits observed in preclinical models translate to humans, and whether sustained TMPRSS6 inhibition may have unintended systemic consequences, remains uncertain.

Genetic studies have identified the *TMPRSS6* locus, and specifically the missense variant rs855791, as a robust determinant of iron homeostasis, with genome-wide significant associations with serum iron, hemoglobin levels, transferrin saturation, and erythrocyte mean cell volume (8,9). Subsequent large-scale genome-wide association studies (GWAS) using magnetic resonance imaging (MRI)-derived liver phenotypes have identified associations between variation at the *TMPRSS6* locus and hepatic iron content, with fine-mapping implicating rs855791 as the likely causal variant (10,11). These findings provide a genetic basis for investigating the consequences of reduced TMPRSS6 activity in humans. However, whether genetically proxied TMPRSS6 inhibition influences hepatic steatosis, fibroinflammatory activity, or clinical liver disease, and whether it carries broader cardiometabolic consequences, remains unclear.

Human genetics offers a powerful tool to prioritize therapeutic targets (12), but its application to some liver outcomes, such as hepatic fat, has been limited by challenges in accurately characterizing liver phenotypes (13,14). Large biobank GWAS have relied on both MRI-derived imaging phenotypes and diagnostic code-based case definitions, and the latter are susceptible to underdiagnosis and misclassification (13). MRI-derived liver phenotypes, including proton density fat fraction (PDFF) as a quantitative measure of hepatic steatosis, and corrected T1 (cT1) as a non-invasive marker of fibroinflammatory activity, provide complementary quantitative measures of hepatic pathology (15–17). Leveraging these phenotypes alongside genetic approaches, such as Mendelian randomization (MR), enables the mechanistic evaluation of the causal pathways underlying hepatic disease and the identification of robust therapeutic targets.

MR is a powerful method that enables the identification of causal effects in humans (18). MR leverages the naturally randomized allocation of genetic variants among the population as instrumental variables, analogous to treatment allocation in a randomized controlled trial. Under certain assumptions, this approach estimates the causal effects of exposures on outcomes, overcoming the limitations of traditional observational studies such as residual confounding and reverse causality (18).

Here, we aimed to evaluate the potential hepatic benefits and systemic safety implications of TMPRSS6 inhibition using human genetic evidence. We leveraged a functional genetic variant as a proxy for reduced TMPRSS6 activity to investigate its associations with MRI-derived hepatic phenotypes, clinical liver disease, and cardiometabolic outcomes. We further integrated biomarker-wide, proteome-wide, and phenome-wide analyses to characterize potential downstream mechanisms and safety liabilities.

## Materials and methods

### Study overview

The study design is summarized in **Fig. 1**. We first identified a biologically plausible genetic instrument to proxy reduced TMPRSS6 activity. Using a two-sample Mendelian randomization framework, we then assessed the associations of genetically proxied TMPRSS6 inhibition with primary hepatic outcomes and secondary cardiometabolic outcomes. To characterize the broader systemic effects of genetically proxied TMPRSS6 inhibition, we performed a phenome-wide scan to identify potential secondary indications and safety liabilities, and biomarker-wide and proteome-wide scans to explore downstream biological pathways and potential markers of target engagement. All analyses were conducted and reported in accordance with the STROBE-MR guidelines (19).

**Fig 1.**
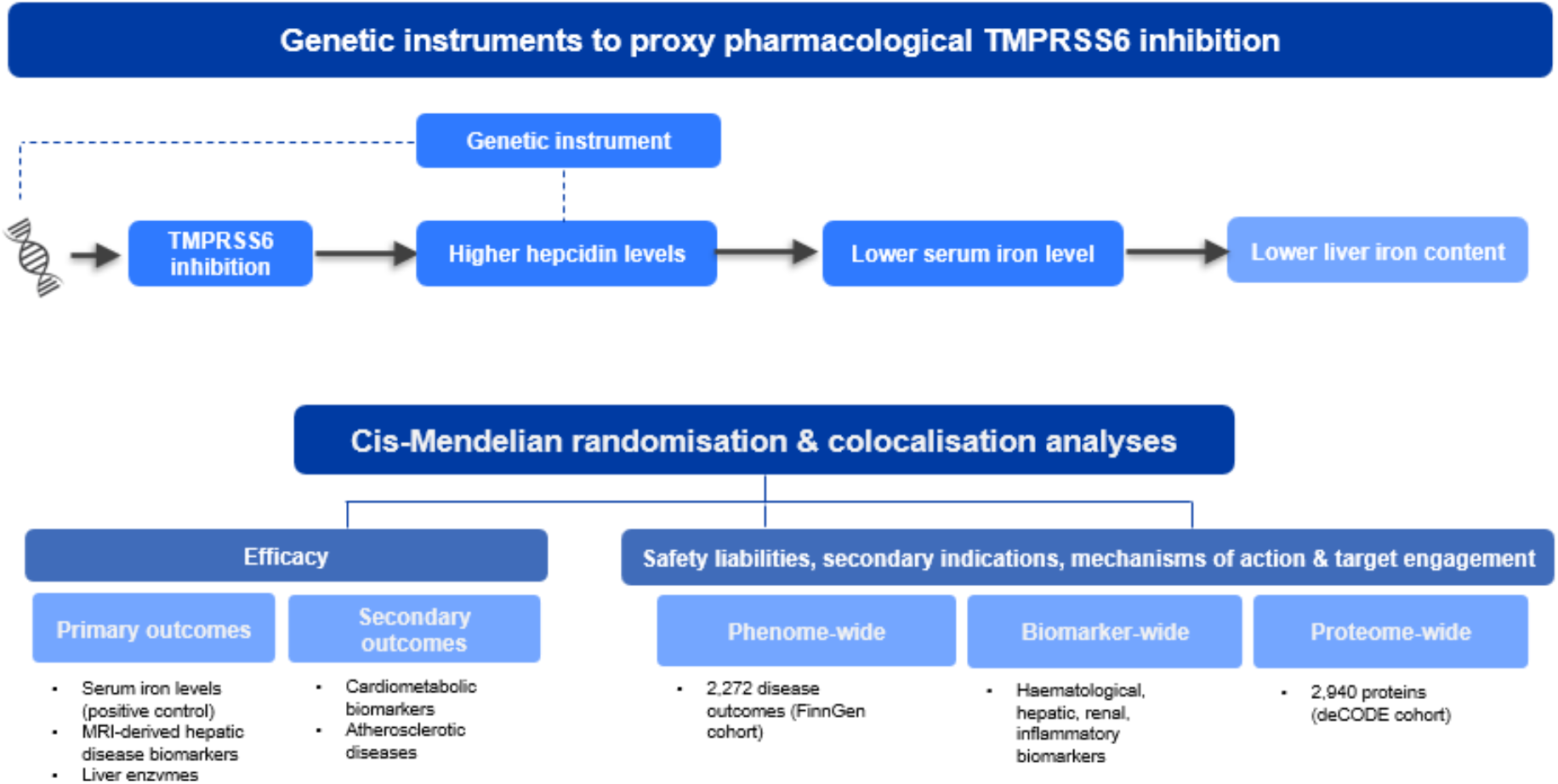
Study Design.

### Instrumental variable selection

To proxy the pharmacological inhibition of TMPRSS6, we used rs855791 (p.Ala736Val), a common functional missense variant in *TMPRSS6* that reduces matriptase-2 serine protease activity, weighted by its effect on circulating hepcidin levels (4). Reduced matriptase-2 activity attenuates suppression of the BMP-SMAD signalling, resulting in increased hepcidin expression. The hepcidin-increasing allele was therefore used to approximate the direction of the downstream biological effects expected from pharmacological TMPRSS6 inhibition.

### Outcome variables and data sources

Details on the GWAS data sources used in our analyses are shown in **Table 1**. GWAS selection was primarily based on sample size and the availability of relevant phenotypes. When available, we used data on associations estimated in European ancestry populations rather than multi-ancestry populations to minimize population stratification.

**Table 1.** Characteristics of genome-wide association study data sources.

| Trait | Unit | N or N<sub>cases</sub>/N<sub>controls</sub> | Ancestry | Cohort(s) | Citation |
|---|---|---|---|---|---|
| Hepcidin | SD of the inverse-normal transformed log-hepcidin (nmol/L) | 91,675 | European | Multi-cohort | Allara et al. 2024 |
| **Positive control** |  |  |  |  |  |
| Serum iron | SD of the IRNT-transformed trait | 127,495 | Trans ancestry | MVP | Verma et al. 2024 |
| **Primary outcomes: markers of hepatic disease** |  |  |  |  |  |
| Liver iron (mg/g dry weight) | Log + IRNT | 37,626 | European | UKB | Meena et al. 2026 |
| cT1; corrected T1 relaxation time (ms) | IRNT | 37,626 | European | UKB | Meena et al. 2026 |
| Proton density fat fraction (PDFF) (%) | Log + IRNT | 37,626 | European | UKB | Meena et al. 2026 |
| Liver volume (L) | SD units of the IRNT-transformed trait | 32,860 | European | UKB | Liu et al. 2021 |
| Alkaline phosphatase (ALP) | Log + IRNT | 437,438 | European | UKB | Pazoki et al. 2021 |
| Alanine aminotransferase (ALT) | Log + IRNT | 437,438 | European | UKB | Pazoki et al. 2021 |
| Gamma-glutamyl transferase (GGT) | Log + IRNT | 437,438 | European | UKB | Pazoki et al. 2021 |
| **Clinical liver outcomes** |  |  |  |  |  |
| Liver cirrhosis | log-odds | 6,421/377,277 | European | FinnGen | Kurki et al. 2023 |
| Nonalcoholic fatty liver disease (NAFLD) | log-odds | 90,408/128,187 | Transancestry | MVP | Vujkovic et al. 2022 |
| Hepatocellular carcinoma (HCC) | log-odds | 3,748/ 1,861,536 | European | Multi-cohort | Ghouse et al. 2025 |
| **Secondary outcomes: cardiometabolic disease** |  |  |  |  |  |
| Body mass index (BMI) | SD units of the IRNT-transformed trait | 806,834 | European | UKB & GIANT | Pulit et al. 2019 |
| Waist-to-hip ratio (WHR) | SD units of the IRNT-transformed trait | 697,734 | European | UKB & GIANT | Pulit et al. 2019 |
| Visceral adipose tissue (VAT) | SD units of the IRNT-transformed trait | 32,860 | European | UKB | Liu et al. 2021 |
| Abdominal subcutaneous adipose tissue (ASAT) | SD units of the IRNT-transformed trait | 32,860 | European | UKB | Liu et al. 2021 |
| Type 2 diabetes (T2D) | log-odds | 180,834/1,159,055 | Transancestry | DIAMANTE | Mahajan et al. 2022 |
| Coronary artery disease (CAD) | log-odds | 181,522/984,168 | European | CARDIoGRAMplusC4D | Aragam et al. 2022 |
| Aortic stenosis (AoStenosis) | log-odds | 86,864/2,766,544 | European | Multi-cohort incl. UKB, FinnGen | Small et al. 2026 |
| Any ischaemic stroke (AIS) | log-odds | 62,100/1,234,808 | European | GIGASTROKE | Mishra et al. 2022 |
| Heart failure | log-odds | 153,174/1,946,349 | European | HERMES | Henry et al. 2025 |
| Atrial fibrillation | log-odds | 181,446/1,468,899 | Transancestry | Multi-cohort | Roselli et al. 2025 |
| **Biomarkers** |  |  |  |  |  |
| *Hematological* |  |  |  |  |  |
| Platelet count (×10<sup>9</sup>/L) | SD units of the IRNT-transformed trait | 542,827 | European | Multi-cohort incl. UKB | Chen et al. 2020 |
| Red blood cell count (×10<sup>12</sup>/L) | SD units of the IRNT-transformed trait | 430,396 | European | Multi-cohort incl. UKB | Chen et al. 2020 |
| Lymphocyte count (×10<sup>9</sup>/L) | SD units of the IRNT-transformed trait | 524,923 | European | Multi-cohort incl. UKB | Chen et al. 2020 |
| Monocyte count (×10<sup>9</sup>/L) | SD units of the IRNT-transformed trait | 521,594 | European | Multi-cohort incl. UKB | Chen et al. 2020 |
| Basophil count (×10<sup>9</sup>/L) | SD units of the IRNT-transformed trait | 474,001 | European | Multi-cohort incl. UKB | Chen et al. 2020 |
| Eosinophil count (×10<sup>9</sup>/L) | SD units of the IRNT-transformed trait | 474,237 | European | Multi-cohort incl. UKB | Chen et al. 2020 |
| Total white blood cell count (×10<sup>9</sup>/L) | SD units of the IRNT-transformed trait | 562,243 | European | Multi-cohort incl. UKB | Chen et al. 2020 |
| Neutrophil count (×10<sup>9</sup>/L) | SD units of the IRNT-transformed trait | 519,288 | European | Multi-cohort incl. UKB | Chen et al. 2020 |
| *Cardiometabolic* |  |  |  |  |  |
| HbA1c (%) | SD units of the IRNT-transformed trait | 144,995 | European | MAGIC | Chen et al. 2021 |
| Fasting glucose (mmol/L) | SD units of the IRNT-transformed trait | 193,036 | European | MAGIC | Chen et al. 2021 |
| Fasting insulin (pmol/L) | Natural log-transformed (ln-pmol/L), then inverse-normal transformed | 146,273 | European | MAGIC | Chen et al. 2021 |
| LDL-c (mmol/L) | SD units of the IRNT-transformed trait | 1,225,532 | European | GLGC | Graham et al. 2021 |
| HDL-c (mmol/L) | SD units of the IRNT-transformed trait | 1,238,209 | European | GLGC | Graham et al. 2021 |
| Triglycerides (mmol/L) | SD units of the IRNT-transformed log-TG | 1,240,000 | European | GLGC | Graham et al. 2021 |
| SBP | mmHg | 1,017,410 | European | UKB + ICBP | Keaton et al. 2024 |
| DBP | mmHg | 1,028,910 | European | UKB + ICBP | Keaton et al. 2024 |
| *Renal* |  |  |  |  |  |
| Creatinine-based eGFR (mL/min/1.73m<sup>2</sup>) | SD change in log-eGFR | 1,650,645 | European | Multi-cohort incl. UKB, MVP, CKDGen | Liu et al. 2025 |
| Cystatin C-based eGFR (mL/min/1.73m<sup>2</sup>) | SD change in log-eGFR | 460,826 | European | CKDGen + UKB | Stanzick et al. 2021 |
| Blood urea nitrogen (mmol/L) | SD change in log-BUN | 370,678 | European | CKDGen + UKB | Wuttke et al. 2019 |
| Urinary albumin-to-creatinine ratio (mg/g) | SD change in log-UACR | 327,616 | European | CKDGen + UKB | Zanetti et al. 2019 |
| *Inflammation* |  |  |  |  |  |
| C-reactive protein (CRP) | SD in log-CRP | 427,367 | European | UKB | Said et al. 2022 |
| **Proteome-wide and phenome-wide analyses** |  |  |  |  |  |
| Plasma proteins (2,940; Olink Explore) | SD of normalised protein expression | 34,557 | European | UKB-PPP | Sun et al. 2023 |
| Disease endpoints (2,272) | log-odds | 377,277 | European | FinnGen | Kurki et al. 2023 |

CKDGen, Chronic Kidney Disease Genetics Consortium; DIAMANTE, DIAbetes Meta-ANalysis of Trans-Ethnic association studies; eGFR, estimated glomerular filtration rate; GIANT, Genetic Investigation of ANthropometric Traits; GLGC, Global Lipids Genetics Consortium; HERMES, Heart Failure Molecular Epidemiology for Therapeutic Targets; ICBP, International Consortium of Blood Pressure; IRNT, inverse rank-normal transformed; MAGIC, Meta-Analyses of Glucose and Insulin-related traits Consortium; MVP, Million Veteran Program; UKB, UK Biobank; UKB-PPP, UK Biobank Pharma Proteomics Project.

We considered serum iron levels (20) as a positive control outcome, to assess whether the selected variant captured the expected downstream effects of TMPRSS6 perturbation on systemic iron homeostasis. As primary outcomes, we investigated markers of hepatic disease, including liver iron (21), cT1 (21), PDFF (21), liver volume (22), alkaline phosphatase (ALP), alanine aminotransferase (ALT), gamma-glutamyl transferase (GGT).

To complement the quantitative MRI phenotypes, we additionally considered clinically defined nonalcoholic fatty liver disease (NAFLD) (23), cirrhosis (24) and hepatocellular carcinoma (HCC) (25). These clinical liver outcomes were analyzed as a separate outcome group.

As secondary outcomes, we considered cardiometabolic traits and disease endpoints, including body mass index (BMI) (26), waist-to-hip ratio (WHR) (26), visceral adipose tissue (VAT) (22), abdominal subcutaneous adipose tissue (ASAT) (22), type 2 diabetes (T2D) (27), coronary artery disease (CAD) (28), aortic stenosis (29), any ischemic stroke (30), heart failure (31) and atrial fibrillation (32).

Phenome-wide analyses included 2,272 disease outcomes derived from FinnGen release 9 (24). Proteome-wide analysis included relative concentrations of 2,940 plasma proteins, measured using the Olink platform (33). Biomarker-wide analysis included a range of hepatic, renal, hematological biomarkers described in detail in **Table 1**.

### Tissue-specific TMPRSS6 expression

Informed by the outcome-level MR results, we next examined whether the observed disease associations were accompanied by evidence of shared genetic signals with tissue-specific *TMPRSS6* expression, using expression quantitative trait locus (eQTL) data from the Genotype Tissue Expression project (GTEx v7), which profiles gene expression across 48 human tissues. We included biologically relevant tissues, including liver and arterial tissues (coronary, tibial, and aortic arteries).

### Statistical Analysis

#### Mendelian randomization analyses

Mendelian randomization rests on three assumptions: the selected genetic instruments must be strongly associated with the exposure, be independent of confounders of the exposure–outcome relationship, and influence the outcome only through the exposure of interest (34).

MR effect estimates were obtained using the Wald ratio estimator. To reflect TMPRSS6 inhibition, MR estimates reflect the change in outcome per standard deviation (SD) increase in genetically proxied circulating hepcidin levels. MR estimates are expressed as beta coefficients with 95% confidence intervals for continuous outcomes and odds ratios (OR) with 95% confidence intervals for binary outcomes. To account for multiple testing, we applied Bonferroni correction separately for primary outcomes (P < 0.007, 7 outcomes), secondary outcomes (P < 0.0045, 10 outcomes), and the biomarker-wide analysis (P < 0.0024, 21 outcomes). For hypothesis-generating analyses (proteome-wide and phenome-wide scans), we used a false-discovery rate (FDR) approach: associations with q < 0.05 were considered significant, and those with unadjusted P < 0.05 but q ≥ 0.05 were considered suggestive associations (35).

#### Colocalization analyses

For primary, secondary, biomarker-wide, or expression outcome where MR analysis indicated a nominally significant result, we performed colocalization as a sensitivity analysis to verify that the two traits were driven by the same variant and shared a common biological mechanism. (36). Colocalization was tested across the *TMPRSS6* gene region (chr22:37,462,936-37,505,753 ± 100 kb, GRCh37) using all SNPs available in both datasets for a given pair. This approach estimates posterior probabilities (PP) across five competing hypotheses: H_0_ (no causal variants in the region), H_1_ (a causal variant for trait 1 only), H_2_ (a causal variant for trait 2 only), H_3_ (separate causal variants for each trait), and H_4_ (a single shared causal variant for both traits, consistent with a true causal relationship). The prior probability of any given SNP being causal for a single trait within a region was set to 10⁻⁴. Evidence for colocalization was classified based on PP.H4 as follows: strong (PP.H4 ≥ 0.80), moderate (0.50 ≤ PP.H4 < 0.80), weak (0.20 ≤ PP.H4 < 0.50), or no colocalization (PP.H4 < 0.20). We additionally computed conditional colocalization [PP.H4/(PP.H4+PP.H4)] to quantify the relative support for a shared versus distinct causal variant, conditioned on at least one causal variant being present for each trait in the region.

#### Mediation analysis

To explore pathways that might underlie the associations between genetically proxied TMPRSS6 inhibition and clinical endpoints identified in primary and secondary analyses, we conducted a two-step (network) MR mediation analysis. Candidate mediators were biomarkers that met two criteria. First, they were associated with genetically proxied TMPRSS6 inhibition in the biomarker-wide MR. Second, they showed at least suggestive evidence of a shared regional genetic signal with the relevant clinical endpoint at the *TMPRSS6* locus in colocalization analyses. The effect of genetically proxied TMPRSS6 inhibition on each mediator (β1) and on each clinical endpoint (total effect, βTE) was obtained from the primary Wald ratio analyses. We estimated the effect of each mediator on the clinical endpoint (β2) using univariable MR with genome-wide instruments for the mediator. Instruments were variants associated with the mediator at P < 5 × 10⁻⁸, clumped at r² < 0.001 within a 10,000 kb window. We excluded variants within ±1 Mb of TMPRSS6, to reduce potential overlap with the target-specific genetic signal used to estimate the exposure–mediator and exposure–outcome associations. The inverse-variance weighted method was the primary estimator. The indirect effect through each mediator was calculated as β1 × β2, and the proportion mediated as β1 × β2 / βTE. The proportion mediated was calculated on the linear scale for continuous outcomes and the log-odds scale for binary outcomes. The 95% confidence intervals were derived using the delta method. Given the assumptions underlying two-step MR mediation and the use of a single variant to proxy TMPRSS6 inhibition, these analyses were considered exploratory.

#### Pathway enrichment analysis

To identify biological pathways enriched among proteins associated with genetically proxied hepcidin, we performed over-representation analysis using clusterProfiler (v4) (37). Proteins reaching FDR significance (q < 0.05) in the proteome-wide MR analysis were tested against Gene Ontology (Biological Process), KEGG, and Reactome pathway databases. The background set was restricted to all 2,940 proteins measured on the Olink panel, rather than the whole genome, to account for the platform’s non-random protein coverage. Enrichment p-values were calculated using a hypergeometric test and corrected for multiple testing using the Benjamini-Hochberg method; pathways with an adjusted p-value < 0.05 were considered significantly enriched.

### Software

All analyses were performed in R version 4.3.2. Mendelian randomization analyses were performed using the ‘MendelianRandomization’ package, and colocalization analyses were performed using the ‘coloc’ package. Pathway enrichment analysis was performed using clusterProfiler, with gene annotation from org.Hs.eg.db and pathway databases queried via KEGG and Reactome (ReactomePA, reactome.db).

## Results

The *TMPRSS6* rs855791 A allele reduces matriptase-2 activity, thereby attenuating suppression of hepcidin expression (β = 0.042, SE = 0.005, P = 1.34 × 10⁻¹⁷; effect allele frequency = 0.436). The selected genetic instrument demonstrated a strong association with circulating hepcidin, with an F statistic of 72.

MR estimates for primary and secondary outcomes are shown in **Fig. 2**. As a positive control outcome, serum iron was strongly inversely associated with genetically proxied TMPRSS6 inhibition (β= −3.65, 95% CI −3.86 to −3.44, P<0.001), consistent with the expected effects of reduced TMPRSS6 activity on iron homeostasis.

**Fig. 2.**
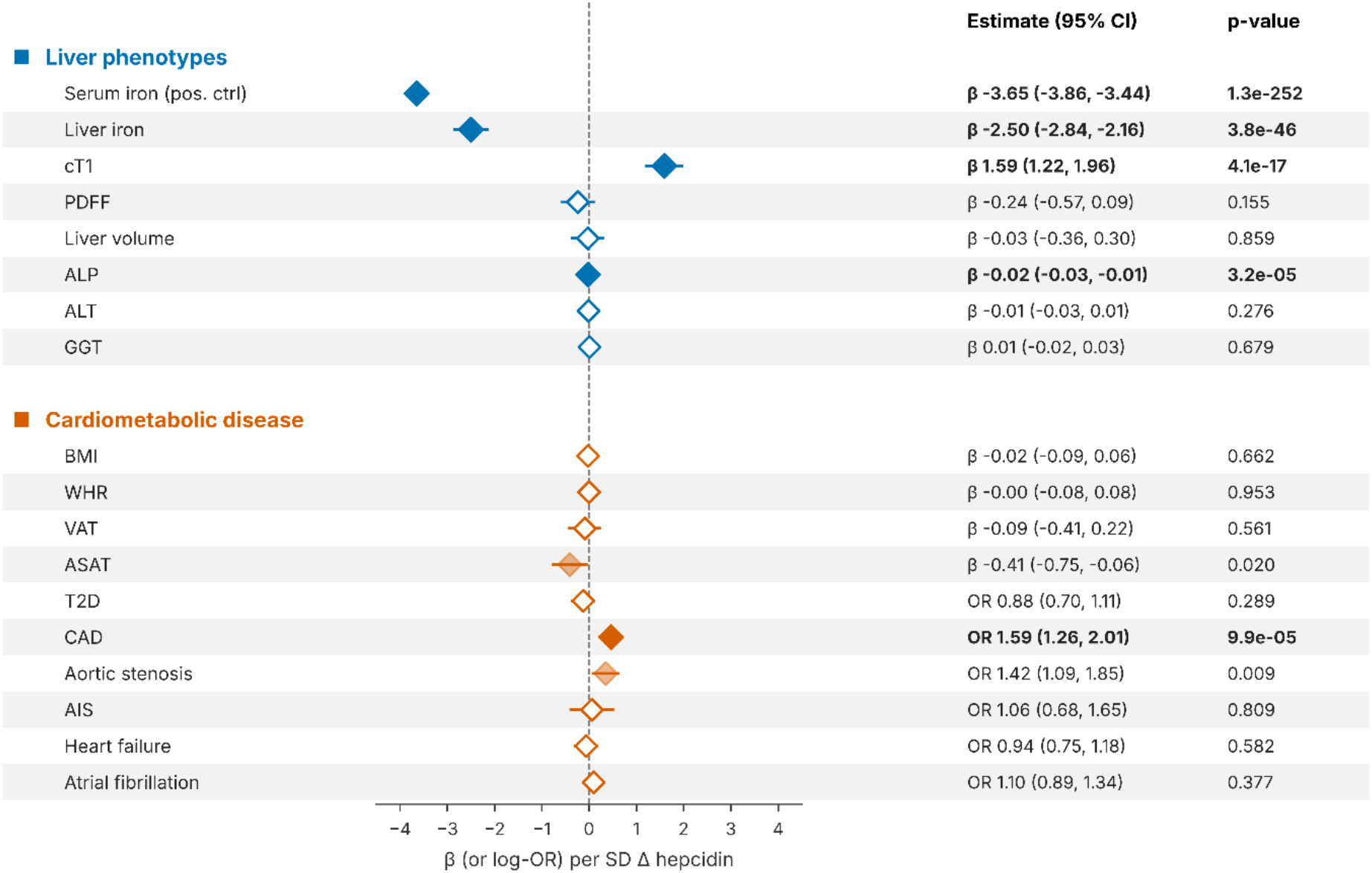
Mendelian randomization estimates for the effect of TMPRSS6 inhibition on primary and secondary outcomes. Liver phenotypes include a positive control (serum iron) and hepatic disease markers. Cardiometabolic disease outcomes include adiposity traits and cardiovascular disease endpoints. Effect estimates represent the SD-unit change in outcome per 1-SD increase in genetically proxied higher hepcidin levels. Liver phenotypes and adiposity traits are reported as β-coefficients (95% CI); disease endpoints (type 2 diabetes, coronary artery disease, aortic stenosis, ischaemic stroke, heart failure, atrial fibrillation) are reported as odds ratios (OR; 95% CI). Filled diamonds indicate associations significant after Bonferroni correction; semi-transparent diamonds indicate nominally significant associations (p < 0.05); open diamonds indicate non-significant associations. The Bonferroni threshold was calculated separately for each outcome group as 0.05 divided by the number of outcomes tested, excluding the positive control (liver phenotypes: α = 0.05/7 = 0.007; cardiometabolic disease: α = 0.05/10 = 0.005).

Among the primary hepatic outcomes, genetically proxied TMPRSS6 inhibition was associated with lower liver iron content (β= −2.50, 95% CI −2.84 to −2.16, P<0.001) and higher cT1 (β=1.59, 95% CI 1.22 to 1.96, P<0.001). There was no evidence of an association with PDFF (β= −0.24, 95% CI −0.57 to 0.09, P=0.15) or liver volume (β= −0.03, 95% CI −0.36 to 0.30, P=0.86). Among hepatic enzymes, genetically proxied TMPRSS6 inhibition was associated with lower ALP levels (β=-0.02, 95% CI −0.03 to −0.01, P<0.001), but there was no evidence of an association with ALT (β=-0.01, 95% CI −0.03 to 0.01, P=0.28) or GGT (β=0.01, 95% CI= −0.02 to 0.03, P=0.68).

For clinically defined liver outcomes **(Supplementary Fig. 1)**, genetically proxied TMPRSS6 inhibition was not associated with NAFLD (OR=1.07, 95% CI 0.78 to 1.46, P=0.69), cirrhosis (OR=0.72, 95% CI 0.15 to 3.42, P=0.68) or hepatocellular carcinoma (OR=0.09, 95% CI 0.00 to 1.96, P=0.12). However, the wide confidence intervals for cirrhosis and hepatocellular carcinoma, precluded definitive conclusions regarding potential protective effects.

For secondary outcomes, there was suggestive evidence of an association between genetically proxied TMPRSS6 inhibition and lower ASAT (β=-0.41, 95% CI= −0.75 to −0.06, P=0.02). There was no evidence of an association with VAT (β=-0.09, 95% CI= −0.41 to 0.22, P=0.56) nor type 2 diabetes (OR=0.88, 95% CI= 0.70 to 1.11, P=0.29).

Genetically proxied TMPRSS6 inhibition was associated with higher risk of several atherosclerotic cardiovascular outcomes, including CAD (OR=1.59, 95% CI= 1.26 to 2.01, P<0.001) and aortic stenosis (OR=1.42, 95% CI= 1.09 to 1.85, P=0.009), with comparable magnitudes of effect. No associations meeting the multiple-testing threshold were observed for ischemic stroke, heart failure or atrial fibrillation.

### Biomarker-wide Mendelian randomization

Biomarker-wide MR estimates are shown in **Fig 3**. Among glucose metabolism traits, genetically proxied TMPRSS6 inhibition was associated with higher HbA1c (β = 0.45, 95% CI 0.39 to 0.51; P = 2.1 × 10⁻⁴⁷), while fasting glucose and fasting insulin showed no association. Among lipid traits, genetically proxied TMPRSS6 inhibition was associated with higher LDL cholesterol (β = 0.22, 95% CI 0.16 to 0.29; P = 2.0 × 10⁻¹¹) and lower triglycerides (β = −0.17, 95% CI −0.24 to −0.11; P = 1.5 × 10⁻⁷).

**Fig 3.**
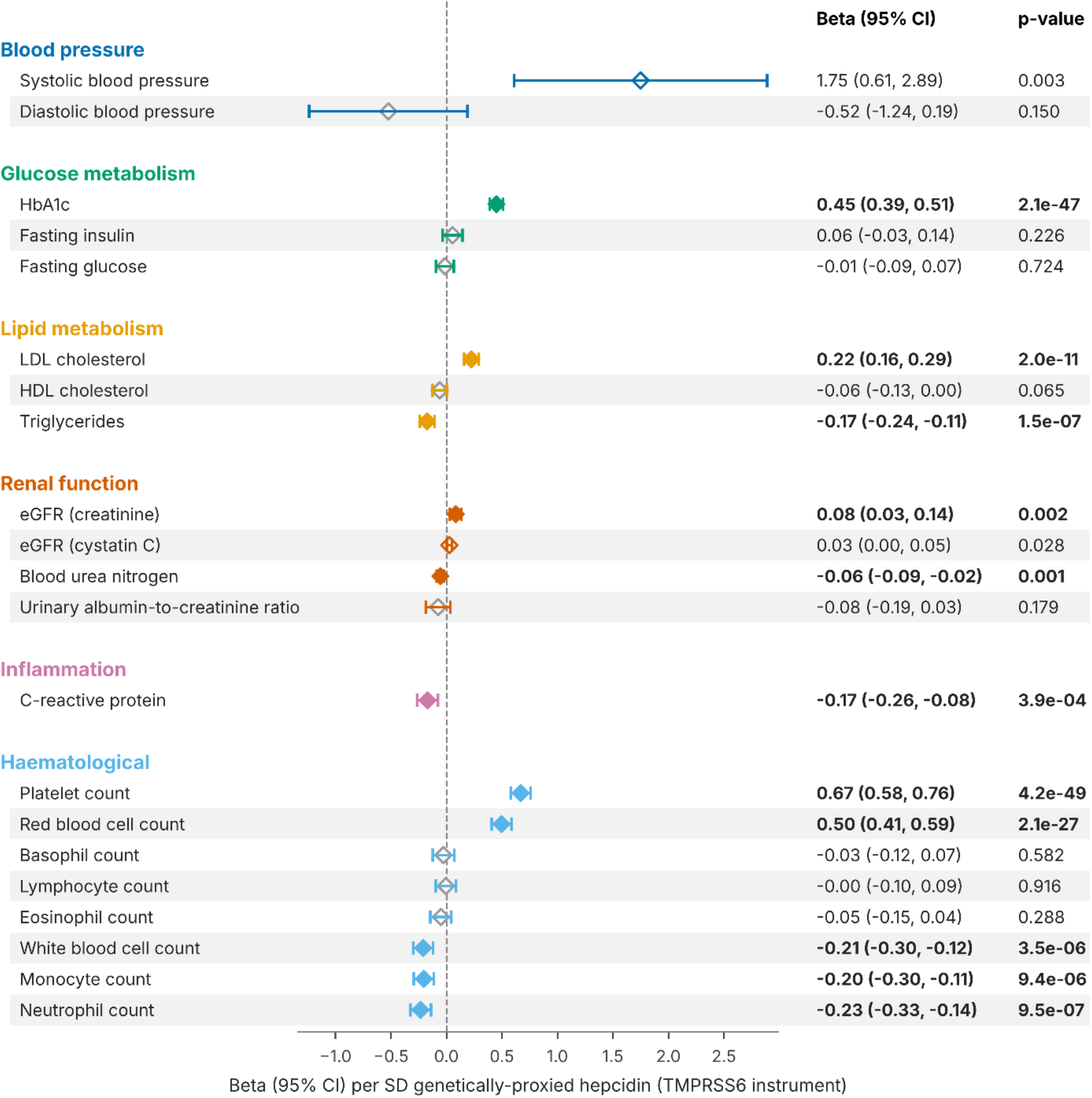
Biomarker-wide Mendelian randomization investigation of TMPRSS6 inhibition. Beta coefficients (95% CI) represent the SD-unit change in each biomarker per 1-SD increase in genetically proxied TMPRSS6 inhibition scaled on increased hepcidin levels. Filled diamonds indicate associations significant after Bonferroni correction (p < 0.0024, accounting for 21 outcomes tested); open colored diamonds indicate nominally significant associations (p < 0.05); open grey diamonds indicate non-significant associations (p ≥ 0.05). Bold text denotes Bonferroni-significant estimates.

Systolic blood pressure was nominally associated with genetically proxied TMPRSS6 inhibition (β = 1.75 mmHg, 95% CI 0.61 to 2.89; P = 0.003), but did not meet the prespecified Bonferroni threshold (P < 0.0024). No association was observed with diastolic blood pressure diastolic blood pressure (β = −0.52 mmHg, 95% CI −1.24 to 0.19; P = 0.150).

Among renal markers, genetically proxied TMPRSS6 inhibition was associated with lower blood urea nitrogen (β = −0.06, 95% CI −0.09 to −0.02; P = 0.001) and higher eGFR estimated using creatinine (β = 0.08, 95% CI 0.03 to 0.14; P = 0.002). eGFR estimated using cystatin C (β = 0.03, 95% CI 0.00 to 0.05; P = 0.028) reached nominal but not Bonferroni significance, while urinary albumin-to-creatinine ratio showed no association (P = 0.179).

Among inflammatory biomarkers, genetically proxied TMPRSS6 inhibition was associated with lower C-reactive protein (β = −0.17, 95% CI −0.26 to −0.08; P = 3.9 × 10⁻⁴).

Among haematological traits, genetically proxied TMPRSS6 inhibition was associated with lower neutrophil count (β = −0.23, 95% CI −0.33 to −0.14; P = 9.5 × 10⁻⁷), monocyte count (β = −0.20, 95% CI −0.30 to −0.11; P = 9.4 × 10⁻⁶), and white blood cell count (β = −0.21, 95% CI −0.30 to −0.12; P = 3.5 × 10⁻⁶), and with higher red blood cell count (β = 0.50, 95% CI 0.41 to 0.59; P = 2.1 × 10⁻²⁷) and platelet count (β = 0.67, 95% CI 0.58 to 0.76; P = 4.2 × 10⁻⁴⁹). Eosinophil, lymphocyte, and basophil counts showed no significant association.

### Colocalization and mechanistic characterization

Cross-trait colocalization results are shown for primary, secondary, and biomarker-wide traits in **Supplementary Fig. 2 and Supplementary Table 3.** Colocalization analysis between liver iron content and cT1 at the *TMPRSS6* gene region yielded a PP.H_4_=99.7%, indicating strong evidence that the associations with liver iron and cT1 share a causal genetic signal.

Colocalization analysis also supported shared regional genetic signals involving hepcidin, HbA1c, platelet count, WBC, neutrophil count, and monocyte count, with PP.H_4_ ≥ 0.99 for all pairwise combinations examined.

CAD risk showed moderate evidence of colocalization with circulating hepcidin at the *TMPRSS6* locus (PP.H_4_ = 0.56; conditional PP = 0.85), consistent with, but not establishing, a shared causal genetic signal **(Fig. 4A, Supplementary Table 3)**. Despite comparable effect sizes and directions in the MR analysis, CAD and calcific aortic stenosis did not show evidence of colocalization (PP.H4 = 0.01; PP.H3 <0.01) **(Fig. 4B, Supplementary Table 3)**.

**Fig 4.**
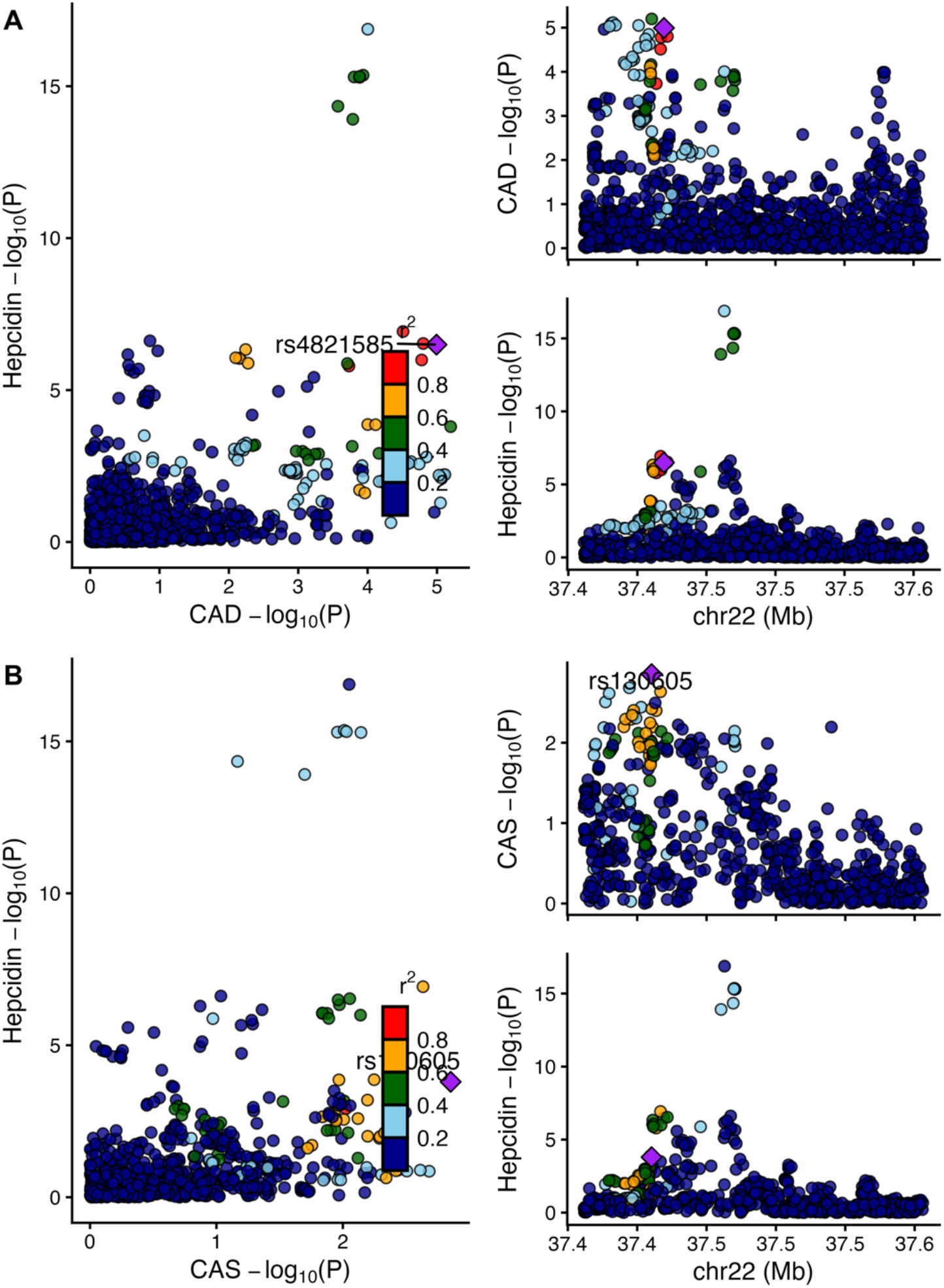
Regional association plots for colocalization analyses at the *TMPRSS6* locus. (A) Regional association plots for genetically proxied hepcidin and coronary artery disease (CAD), showing weak evidence of colocalization (PP.H_4_ = 0.56; conditional PP = 0.85). (B) Regional association plots for CAD and calcific aortic stenosis, showing little evidence of colocalization (PP.H4 = 0.01; PP.H3<0.01) despite comparable directions and magnitudes of association in the Mendelian randomization analysis.

CAD risk also showed weak-to-moderate support for colocalization with HbA1c, platelet count, and LDL-c (PP.H_4_ = 0.48-0.57), but showed limited support for colocalization with CRP (PP.H_4_ <0.01) or systolic blood pressure (PP.H_4_ <0.01). Total cholesterol and LDL-c showed strong evidence of colocalization with hepcidin and HbA1c (PP.H_4_ = 0.996-0.999), supporting shared regional genetic signals across iron-related, glycaemic, and lipid traits.

We next tested whether CAD risk could be explained by tissue-specific TMPRSS6 expression, using cis-eQTLs from three arterial tissues (aorta, coronary artery, and tibial artery). No evidence of colocalization was observed (all PP.H4 < 0.001; Supplementary Table 3a). However, none of the artery eQTL datasets contained genome-wide significant cis-eQTLs at the *TMPRSS6* locus.

### Two-step mediation analysis

We included LDL-C and platelet count in the mediation analysis because both biomarkers were associated with genetically proxied TMPRSS6 inhibition and showed at least weak to moderate evidence of colocalization with CAD at the *TMPRSS6* locus. Higher genetically predicted LDL-C was associated with greater CAD risk (OR 1.55 per SD, 95% CI 1.46 to 1.64, P = 6.4 × 10⁻⁴⁶), whereas genetically predicted platelet count was not (OR 0.99 per SD, 95% CI 0.95 to 1.02, P = 0.53) **(Fig. 5)**. Results were consistent across the weighted median and MR-Egger estimators, with no evidence of directional pleiotropy (MR-Egger intercept P = 0.23 for LDL-C and P = 0.55 for platelet count). In exploratory two-step MR mediation analysis, LDL-C accounted for an estimated 21.2% (95% CI 8.5%–33.9%) of the association between genetically proxied TMPRSS6 inhibition and CAD. In contrast, there was no evidence of mediation through platelet count (−1.7%, 95% CI −6.9% to 3.6%). These estimates should be interpreted in the context of the assumptions underlying genetic mediation analysis.

**Fig 5.**
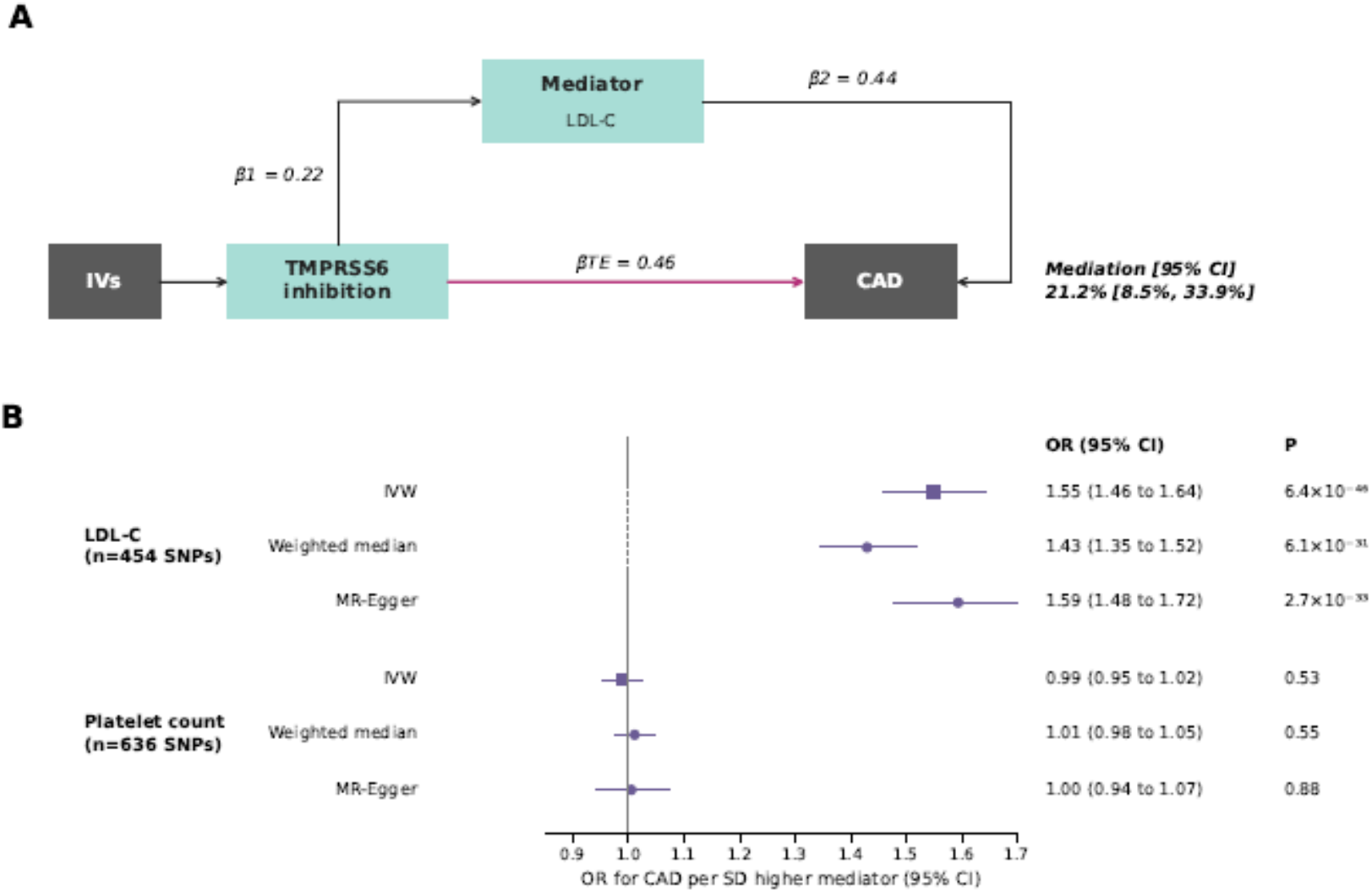
Mediation of the effect of genetically proxied TMPRSS6 inhibition on coronary artery disease. (A) Proportion of the effect of TMPRSS6 inhibition on CAD mediated through LDL-C. β1, effect of genetically proxied TMPRSS6 inhibition (per SD higher hepcidin levels) on LDL-C; β2, effect of LDL-C on CAD (log OR per SD); βTE, total effect of TMPRSS6 inhibition on CAD (log OR). Proportion mediated was estimated as β1 × β2/βTE, with 95% confidence intervals derived using the delta method. (B) Genome-wide MR estimates for the effects of LDL-C and platelet count on CAD, using inverse-variance weighted (squares), weighted median and MR-Egger (circles) estimators.

### Proteome-wide Mendelian randomization

Of the 2,940 tested plasma proteins, 30 were associated with genetically predicted TMPRSS6 inhibition **(Fig 6, Supplementary Table 4)**. The strongest associations were with iron-transport and erythropoiesis-related proteins: TFRC (β = 3.27; P = 9.9 × 10^-134^), AHSP (β = −1.82; P = 6.6 × 10^-39^), EPO (β = 1.82; P = 3.5 × 10^-35^), MELTF (β = −1.43; P = 7.0 × 10^-28^), and CSF2RB (β = −1.35; P = 1.7 × 10⁻¹⁹). Several heme-biosynthesis proteins were also associated, including CPOX (β = −1.46; P = 1.8 × 10⁻²²), HMBS (β = −0.97; P = 7.7 × 10⁻¹⁴), and ARG1 (β = 0.55; P = 1.5 × 10⁻¹⁰). Directionally, proteins involved in iron acquisition and transport (TFRC, EPO, TF) increased with genetically proxied TMPRSS6 inhibition, while proteins involved in heme synthesis (HMBS, CPOX, ARG1) decreased, consistent with hepcidin-mediated restriction of iron availability for erythropoiesis.

**Fig 6.**
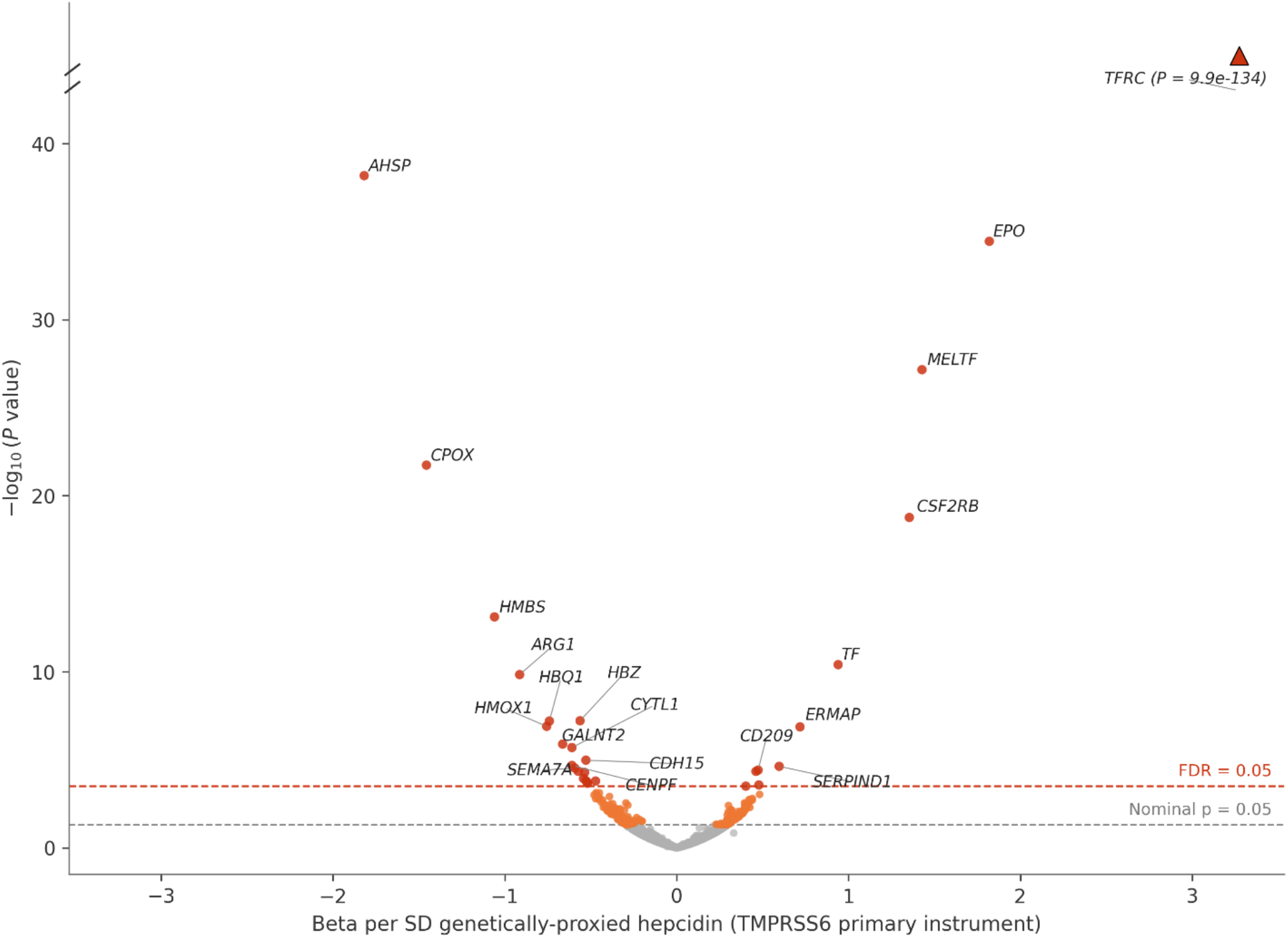
Proteome-wide Mendelian randomization estimates for the effect of TMPRSS6 inhibition on plasma protein levels. Betas represent the SD change in relative protein abundance per SD increase in genetically proxied TMPRSS6 inhibition, scaled on increased hepcidin levels. Points are coloured by significance tier: FDR-significant (red), nominally significant (orange), and not significant (grey). Proteins are labeled by their gene symbol. The grey dashed line represents nominal significance (P < 0.05); the red dashed line represents the FDR-adjusted significance threshold (FDR < 0.05, corresponding to raw P < 3.1 × 10⁻⁴). TFRC’s estimate is truncated at the axis limit (▴) due to its extreme significance (P = 9.9 × 10⁻¹³⁴); its true value is annotated on the plot.

Over-representation analysis (GO, KEGG, Reactome; background restricted to the Olink panel) showed that the 30 FDR-significant proteins clustered in three related processes: iron transport (MELTF, TF, TFRC), heme/porphyrin metabolism (CPOX, HMBS, HMOX1), and erythrocyte/myeloid homeostasis (AHSP, EPO, HBQ1, HBZ, HMOX1), with additional enrichment for HIF-1 signaling and ferroptosis **(Supplementary Table 5)**. This is consistent with a proteomic signature centered on iron acquisition, heme synthesis, and erythropoiesis.

### Phenome-wide Mendelian randomization

Genetically proxied TMPRSS6 inhibition was not associated with any of the 2,272 PheWAS endpoints at the FDR-significant threshold **(Supplementary Table 6).** The strongest signal was observed for anemia-related phenotypes, consistent with the established role of TMPRSS6 in iron homeostasis and validating genetic instrument selection.

## Discussion

In this large-scale Mendelian randomization study, we leveraged human genetic variation at the *TMPRSS6* locus to investigate the potential hepatic benefits and systemic safety implications of reduced TMPRSS6 activity across hepatic and cardiometabolic outcomes. Our analyses yield three principal findings with potential implications for therapeutic development. First, genetically proxied TMPRSS6 inhibition was associated with lower liver iron content but paradoxically higher cT1, with no clear association with PDFF or liver volume, suggesting that hepatic consequences of TMPRSS6 inhibition are specific to iron handling rather than broad metabolic perturbation. Second, genetically proxied TMPRSS6 inhibition was associated with increased risk of coronary artery disease (OR = 1.59), alongside a nominal association with aortic stenosis (OR = 1.42). Third, genetically proxied TMPRSS6 inhibition was associated with higher LDL-C, which exploratory mediation analysis identified as a potential contributor to the CAD association.

### Hepatic effects: iron restriction without evidence of steatosis reduction

The genetic evidence provided by our main analysis suggests that TMPRSS6 inhibition reduces liver iron content, consistent with its established mechanism: impaired matriptase-2 serine protease activity elevates hepcidin, which promotes ferroportin internalization and degradation on enterocytes and reticuloendothelial macrophages, restricting dietary iron absorption and recycling (3,38). Alongside the reduction in liver iron content, genetically proxied TMPRSS6 inhibition was associated with higher cT1, a quantitative multiparametric MRI marker used as a non-invasive index of hepatic fibroinflammatory activity (21). These associations were directionally opposite, yet colocalization analysis provided strong evidence that the associations share a causal genetic signal at the *TMPRSS6* locus. This divergence in effects may reflect the influence of hepatic iron on the cT1 measurement itself rather than a true increase in fibroinflammatory activity. Iron is paramagnetic and shortens tissue T1 relaxation time; consequently, iron accumulation can suppress measured T1 values, whereas a reduction in hepatic iron may increase cT1 through removal of this T1-shortening effect (39). Thus, the higher cT1 associated with genetically proxied TMPRSS6 inhibition may, at least in part, represent an imaging consequence of reduced hepatic iron. In our complementary analyses examining clinically defined liver outcomes as orthogonal endpoints, genetically proxied TMPRSS6 inhibition was not significantly associated with NAFLD, cirrhosis or hepatocellular carcinoma. If the higher cT1 reflected a true increase in fibroinflammatory activity, a concordant increase in the risk of advanced liver disease would have been expected. The point estimates for cirrhosis and hepatocellular carcinoma were in the direction of lower risk, opposite to that implied by the cT1 association. This is more consistent with the cT1 association being, at least in part, an imaging consequence of reduced hepatic iron. Nevertheless, these estimates were imprecise and cannot exclude clinically relevant effects in either direction.

Notably, no association was observed with hepatic steatosis as measured by PDFF, NAFLD nor with liver volume. This contrasts with the preclinical findings of Pettinato et al., in which GalNAc-conjugated antisense oligonucleotide-mediated Tmprss6 knockdown reduced both steatohepatitis and fibrosis in murine models of MASLD (6). Several factors may account for this apparent discrepancy. The anti-steatotic effects observed in preclinical models may not translate directly to humans, may depend on disease context or baseline iron burden, or may require a degree of target inhibition and consequent iron restriction that is not adequately recapitulated by lifelong exposure to common genetic variants. The absence of a detectable association with PDFF in the present study does not exclude an effect of pharmacological TMPRSS6 inhibition on hepatic steatosis. This is consistent with prior evidence from human genetic studies showing that the TMPRSS6 p.Ala736Val variant independently associates with hepatic iron accumulation in NAFLD, with the higher-hepcidin allele conferring lower hepatic iron stores (40), and with transcriptomic data demonstrating upregulated hepatic TMPRSS6 expression in NASH, suggesting a reactive counter-regulatory response to hepatic inflammation (41).

### Cardiovascular safety and potential mechanisms

A key finding of this study was the association between genetically proxied TMPRSS6 inhibition and increased risk of CAD. This observation is particularly relevant given the complex and historically debated relationship between systemic iron status and cardiovascular disease. Epidemiological studies have reported associations between circulating iron biomarkers and coronary outcomes (42), while human genetic studies have provided evidence that perturbation of systemic iron status may influence atherosclerotic cardiovascular risk (43–45). Our findings extend this literature by suggesting that genetically mediated perturbation of the TMPRSS6-hepcidin axis may have cardiovascular consequences that warrant consideration during therapeutic development.

Hepcidin sits at the intersection of iron regulation and inflammatory signaling, with inflammatory pathways, particularly IL-6/JAK/STAT signaling, capable of increasing hepcidin expression independently of systemic iron stores (46). Conversely, changes in iron availability can influence oxidative stress, cellular metabolism, erythropoiesis and vascular biology. TMPRSS6 therefore operates within a regulatory network in which iron, inflammatory and cardiometabolic pathways intersect. However, our colocalization analyses provide little support for systemic inflammation as the principal explanation for the observed CAD association. Although genetically proxied TMPRSS6 inhibition was associated with lower CRP, the CRP signal did not colocalize with CAD at the TMPRSS6 locus. Similarly, systolic blood pressure showed only a nominal association in the biomarker-wide analysis and did not colocalize with CAD.

In contrast, lipid metabolism emerged as a potential pathway linking TMPRSS6 inhibition to coronary risk. Genetically proxied TMPRSS6 inhibition was associated with higher LDL-C, although evidence of a shared causal variant between LDL-C and CAD at the TMPRSS6 locus was moderate. Mediation analyses suggested that LDL-C may account for around one fifth of the CAD association, with uncertainty around the estimated mediated proportion. This finding should be regarded as hypothesis-generating, given the modest colocalization evidence and the reliance of the CAD estimate on a single variant. The remaining association may involve other pathways, although the mediation estimate does not establish their presence or relative contributions. Platelet count was also associated with TMPRSS6 inhibition and showed moderate evidence of a shared signal with CAD, but genetically predicted platelet count was not associated with CAD, and we found no evidence of mediation. This suggests that the platelet signal reflects shared genetic architecture at the locus, rather than a causal pathway to CAD. The mechanisms underlying the CAD association therefore remain largely unexplained.

We additionally investigated whether the CAD association could reflect altered TMPRSS6 expression within the vasculature. No evidence of colocalization was observed between CAD and TMPRSS6 expression in aortic, coronary or tibial artery tissue. Interpretation of these analyses is limited, however, by the absence of genome-wide significant cis-eQTLs for TMPRSS6 in the available arterial datasets, which may reflect insufficient statistical power or low and context-dependent expression of TMPRSS6 in these tissues. Larger tissue-specific eQTL datasets and cell-type-resolved functional studies will be required to determine whether local vascular effects contribute to the observed association.

### Systemic metabolic and haematological consequences of TMPRSS6 inhibition

The association with higher HbA1c, in the absence of corresponding effects on fasting glucose or insulin, may reflect alterations in erythrocyte biology rather than impaired glycemic control. HbA1c is influenced by erythrocyte lifespan and turnover independently of circulating glucose, an interpretation of particular relevance given the pronounced hematological effects of genetically proxied TMPRSS6 inhibition, including higher red blood cell and platelet counts. Similarly, the association with higher creatinine-based eGFR suggest potential effects on renal biomarkers, although these findings should not be interpreted as evidence of improved kidney function. Iron homeostasis is closely linked to renal physiology through the hepcidin-ferroportin axis, which is frequently dysregulated in chronic kidney disease (47), and systemic changes in iron availability may influence renal and hematological biomarkers through several pathways. The discordance between creatinine- and cystatin C-based eGFR associations further warrants caution in attributing these findings to changes in kidney function.

### Clinical implications

These findings have potential implications for the clinical development of TMPRSS6-directed therapies. In trials targeting systemic iron overload, cardiovascular outcomes and lipid profiles should be evaluated alongside established pharmacodynamic measures of iron restriction. This consideration may be particularly important for potential liver indications, where treatment may be prolonged and patients with MASLD frequently have substantial underlying cardiometabolic risk. Lifelong genetically proxied perturbation is not equivalent to pharmacological inhibition over a defined treatment period, and our results therefore do not establish that therapeutic TMPRSS6 inhibition will increase cardiovascular events. Nevertheless, the convergence of the MR and colocalization findings identifies CAD as a potential on-target safety liability that warrants prospective investigation. Monitoring LDL-c and other atherogenic lipid measures during early-phase trials may help determine whether the genetic lipid signal is recapitulated pharmacologically and whether cardiovascular risk mitigation strategies are required during longer-term treatment.

### Strengths and limitations

This study has several important strengths. The use of a functional missense variant in TMPRSS6 provides a biologically informed proxy for reduced matriptase-2 activity, strengthening attribution of the genetic perturbation to the therapeutic target. The use of quantitative MRI-derived liver phenotypes reduces phenotypic misclassification relative to diagnostic code-based definitions and enables mechanistic dissection of steatosis and fibro inflammation as separable endpoints. The breadth of the phenome-wide and proteome-wide screens provides a comprehensive pharmacological landscape that simultaneously informs therapeutic positioning and safety surveillance. Colocalization analyses allow us to assess whether regional associations are consistent with shared genetic signals rather than distinct variants in linkage disequilibrium.

Several limitations merit acknowledgment. As with all MR analyses, the exclusion restriction assumption, that genetic instruments act on outcomes exclusively through TMPRSS6 inhibition, cannot be formally verified, and pleiotropy cannot be entirely excluded. Our analysis relied on a single missense variant and the Wald ratio estimator. Although the functional annotation of rs855791 and its expected association with serum iron support biological relevance, they do not establish the exclusion restriction assumption. We used colocalization to help distinguish shared regional signals from linkage disequilibrium confounding, but we cannot exclude horizontal pleiotropy or demonstrate that all observed associations reproduce the effects of pharmacological TMPRSS6 inhibition. Analyses are conducted predominantly in individuals of European ancestry, which may limit the generalizability of findings to other populations. The genetic instruments capture lifelong, germline variation and may not fully recapitulate the pharmacodynamics of a potent pharmacological inhibitor, particularly with respect to dose magnitude and temporal dynamics.. Finally, the proteome-wide analyses require replication and mechanistic follow-up to distinguish robust downstream biological effects from associations that may reflect pleiotropy or other sources of bias.

## Conclusions

This study provides human genetic evidence to support an association for genetically proxied TMPRSS6 inhibition with lower hepatic iron content, but does not provide clear evidence of reduced hepatic steatosis or clinical liver disease risk, despite encouraging preclinical findings. The accompanying increase in cT1 requires cautious interpretation in the context of altered iron homeostasis and does not establish worsening hepatic fibroinflammatory activity. The association with increased CAD risk identifies a potential cardiovascular safety signal, with LDL-C representing a candidate pathway requiring further investigation. Further clinical studies are needed to assess hepatic efficacy and cardiovascular safety, alongside prospective monitoring of iron, hematological and lipid parameters.

## Abbreviations

ALP: Alkaline phosphatase
ALT: Alanine aminotransferase
ASAT: Abdominal subcutaneous adipose tissue
BMI: Body mass index
BMP: Bone morphogenetic protein
CAD: Coronary artery disease
CI: Confidence interval
CPOX: Coproporphyrinogen oxidase
CRP: C-reactive protein
cT1: Corrected T1
eGFR: Estimated glomerular filtration rate
EPO: Erythropoietin
eQTL: Expression quantitative trait locus
FDR: False discovery rate
GalNAc: N-acetylgalactosamine
GGT: Gamma-glutamyl transferase
GO: Gene Ontology
GTEx: Genotype-Tissue Expression
GWAS: Genome-wide association study
HbA1c: Glycated haemoglobin
HIF-1: Hypoxia-inducible factor 1
KEGG: Kyoto Encyclopedia of Genes and Genomes
LDL-C: Low-density lipoprotein cholesterol
MASLD: Metabolic dysfunction-associated steatotic liver disease
MASH: Metabolic dysfunction-associated steatohepatitis
MR: Mendelian randomization
MRI: Magnetic resonance imaging
NAFLD: Non-alcoholic fatty liver disease
OR: Odds ratio
PDFF: Proton density fat fraction
PheWAS: Phenome-wide association study
PP: Posterior probability
RBC: Red blood cell
SD: Standard deviation
SE: Standard error
SNP: Single-nucleotide polymorphism
T2D: Type 2 diabetes
TF: Transferrin
TFRC: Transferrin receptor 1
TMPRSS6: Transmembrane protease serine 6
UACR: Urinary albumin-to-creatinine ratio
VAT: Visceral adipose tissue
WBC: White blood cell
WHR: Waist-to-hip ratio

## Data availability statement

This study used publicly available summary statistics from genome-wide association studies, cited in the Methods.

## References

1. Devarbhavi H, Asrani SK, Arab JP, Nartey YA, Pose E, Kamath PS. Global burden of liver disease: 2023 update. J Hepatol. 2023 Aug;79(2):516–37. doi:10.1016/j.jhep.2023.03.017 PubMed PMID: 36990226.

2. Teng ML, Ng CH, Huang DQ, Chan KE, Tan DJ, Lim WH, et al. Global incidence and prevalence of nonalcoholic fatty liver disease. Clin Mol Hepatol. 2023 Feb;29(Suppl):S32–42. doi:10.3350/cmh.2022.0365 PubMed PMID: 36517002; PubMed Central PMCID: PMC10029957.

3. Folgueras AR, de Lara FM, Pendás AM, Garabaya C, Rodríguez F, Astudillo A, et al. Membrane-bound serine protease matriptase-2 (Tmprss6) is an essential regulator of iron homeostasis. Blood. 2008 Sep 15;112(6):2539–45. doi:10.1182/blood-2008-04-149773 PubMed PMID: 18523150.

4. Enns CA, Weiskopf T, Zhang RH, Wu J, Jue S, Kawaguchi M, et al. Matriptase-2 regulates iron homeostasis primarily by setting the basal levels of hepatic hepcidin expression through a nonproteolytic mechanism. J Biol Chem. 2023 Oct;299(10):105238. doi:10.1016/j.jbc.2023.105238 PubMed PMID: 37690687; PubMed Central PMCID: PMC10551898.

5. Finberg KE, Whittlesey RL, Fleming MD, Andrews NC. Down-regulation of Bmp/Smad signaling by Tmprss6 is required for maintenance of systemic iron homeostasis. Blood. 2010 May 6;115(18):3817–26. doi:10.1182/blood-2009-05-224808 PubMed PMID: 20200349; PubMed Central PMCID: PMC2865872.

6. Pettinato M, Furiosi V, Carleo R, Bavuso Volpe L, Guo S, Mannella V, et al. Targeting the Liver Serine Protease TMPRSS6 Ameliorates Steatosis and Attenuates Fibrosis in Experimental MASLD. Liver Int. 2025 Jul;45(7):e70163. doi:10.1111/liv.70163 PubMed PMID: 40501083; PubMed Central PMCID: PMC12159589.

7. Rottey S, Singh N, Labriola-Tompkins E, Lob HE, Mohammadi K, Hassan H, et al. Single Ascending Doses of REGN7999, a Monoclonal Antibody Inhibitor of TMPRSS6, Increase Serum Hepcidin and Cause Deep, Sustained Reductions in Serum Iron in Healthy Human Volunteers. Blood. 2023 Nov 2;142(Supplement 1):3841–3841. doi:10.1182/blood-2023-187643

8. Chambers JC, Zhang W, Li Y, Sehmi J, Wass MN, Zabaneh D, et al. Genome-wide association study identifies variants in TMPRSS6 associated with hemoglobin levels. Nat Genet. 2009 Nov;41(11):1170–2. doi:10.1038/ng.462 PubMed PMID: 19820698; PubMed Central PMCID: PMC3178047.

9. Benyamin B, Ferreira MAR, Willemsen G, Gordon S, Middelberg RPS, McEvoy BP, et al. Common variants in TMPRSS6 are associated with iron status and erythrocyte volume. Nat Genet. 2009 Nov;41(11):1173–5. doi:10.1038/ng.456 PubMed PMID: 19820699; PubMed Central PMCID: PMC3135421.

10. Wilman HR, Parisinos CA, Atabaki-Pasdar N, Kelly M, Thomas EL, Neubauer S, et al. Genetic studies of abdominal MRI data identify genes regulating hepcidin as major determinants of liver iron concentration. Journal of Hepatology. 2019 Sep;71(3):594–602. doi:10.1016/j.jhep.2019.05.032

11. Meena D, Pansini M, Fichera A, Huang J, Ahmed A, Dehghan A, et al. Unravelling genetic susceptibility and causal factors in liver health using MRI quantification of inflammation, fat and iron in the liver. Hum Genomics. 2026 Feb 28;20(1):58. doi:10.1186/s40246-026-00913-2 PubMed PMID: 41761370; PubMed Central PMCID: PMC13019979.

12. Nelson MR, Tipney H, Painter JL, Shen J, Nicoletti P, Shen Y, et al. The support of human genetic evidence for approved drug indications. Nat Genet. 2015 Aug;47(8):856–60. doi:10.1038/ng.3314

13. Chen VL, Kuppa A, Oliveri A, Chen Y, Ponnandy P, Patel PB, et al. Human genetics of metabolic dysfunction–associated steatotic liver disease: from variants to cause to precision treatment. Journal of Clinical Investigation. 2025 Apr 1;135(7):e186424. doi:10.1172/JCI186424

14. Bilson J, Yaghootkar H. Genetics of MASLD: a diabetes perspective. Diabetologia. 2026 Nov;69(11):3105–20. doi:10.1007/s00125-026-06759-6 PubMed PMID: 42257871; PubMed Central PMCID: PMC13627228.

15. Parisinos CA, Wilman HR, Thomas EL, Kelly M, Nicholls RC, McGonigle J, et al. Genome-wide and Mendelian randomisation studies of liver MRI yield insights into the pathogenesis of steatohepatitis. J Hepatol. 2020 Aug;73(2):241–51. doi:10.1016/j.jhep.2020.03.032 PubMed PMID: 32247823; PubMed Central PMCID: PMC7372222.

16. Ahmed A, Cule M, Naz A, Thanaj M, Sorokin EP, Odoemelam CS, et al. MRI-Based Genetic Studies Reveal Specific Genetic Variants and Disease Risks Associated With Fat Distribution Across Anatomical Sites. J Obes. 2025;2025:7792701. doi:10.1155/jobe/7792701 PubMed PMID: 40922984; PubMed Central PMCID: PMC12414625.

17. Meena D, Pansini M, Fichera A, Huang J, Ahmed A, Dehghan A, et al. Unravelling genetic susceptibility and causal factors in liver health using MRI quantification of inflammation, fat and iron in the liver. Hum Genomics. 2026 Feb 28;20(1):58. doi:10.1186/s40246-026-00913-2 PubMed PMID: 41761370; PubMed Central PMCID: PMC13019979.

18. Davies NM, Holmes MV, Davey Smith G. Reading Mendelian randomisation studies: a guide, glossary, and checklist for clinicians. BMJ. 2018 Jul 12;k601. doi:10.1136/bmj.k601

19. Skrivankova VW, Richmond RC, Woolf BAR, Davies NM, Swanson SA, VanderWeele TJ, et al. Strengthening the reporting of observational studies in epidemiology using mendelian randomisation (STROBE-MR): explanation and elaboration. BMJ. 2021 Oct 26;375:n2233. doi:10.1136/bmj.n2233 PubMed PMID: 34702754; PubMed Central PMCID: PMC8546498.

20. Verma A, Huffman JE, Rodriguez A, Conery M, Liu M, Ho YL, et al. Diversity and scale: Genetic architecture of 2068 traits in the VA Million Veteran Program. Science. 2024 Jul 19;385(6706):eadj1182. doi:10.1126/science.adj1182

21. Meena D, Pansini M, Fichera A, Huang J, Ahmed A, Dehghan A, et al. Unravelling genetic susceptibility and causal factors in liver health using MRI quantification of inflammation, fat and iron in the liver. Hum Genomics. 2026 Feb 28;20(1):58. doi:10.1186/s40246-026-00913-2 PubMed PMID: 41761370; PubMed Central PMCID: PMC13019979.

22. Liu Y, Basty N, Whitcher B, Bell JD, Sorokin EP, Van Bruggen N, et al. Genetic architecture of 11 organ traits derived from abdominal MRI using deep learning. eLife. 2021 Jun 15;10:e65554. doi:10.7554/eLife.65554

23. Vujkovic M, Ramdas S, Lorenz KM, Guo X, Darlay R, Cordell HJ, et al. A multiancestry genome-wide association study of unexplained chronic ALT elevation as a proxy for nonalcoholic fatty liver disease with histological and radiological validation. Nat Genet. 2022 Jun;54(6):761–71. doi:10.1038/s41588-022-01078-z

24. Kurki MI, Karjalainen J, Palta P, Sipilä TP, Kristiansson K, Donner KM, et al. FinnGen provides genetic insights from a well-phenotyped isolated population. Nature. 2023 Jan 19;613(7944):508–18. doi:10.1038/s41586-022-05473-8

25. Ghouse J, Gellert-Kristensen H, O’Rourke CJ, Seidelin AS, Thorleifsson G, Sveinbjörnsson G, et al. Genome-wide meta-analysis identifies nine loci associated with higher risk of hepatocellular carcinoma development. JHEP Reports. 2025 Sep;7(9):101485. doi:10.1016/j.jhepr.2025.101485

26. Pulit SL, Stoneman C, Morris AP, Wood AR, Glastonbury CA, Tyrrell J, et al. Meta-analysis of genome-wide association studies for body fat distribution in 694 649 individuals of European ancestry. Human Molecular Genetics. 2019 Jan 1;28(1):166–74. doi:10.1093/hmg/ddy327

27. Mahajan A, Spracklen CN, Zhang W, Ng MCY, Petty LE, Kitajima H, et al. Multi-ancestry genetic study of type 2 diabetes highlights the power of diverse populations for discovery and translation. Nat Genet. 2022 May;54(5):560–72. doi:10.1038/s41588-022-01058-3 PubMed PMID: 35551307; PubMed Central PMCID: PMC9179018.

28. Aragam KG, Jiang T, Goel A, Kanoni S, Wolford BN, Atri DS, et al. Discovery and systematic characterization of risk variants and genes for coronary artery disease in over a million participants. Nat Genet. 2022 Dec;54(12):1803–15. doi:10.1038/s41588-022-01233-6

29. Small AM, Yang TY, Itoh S, Thériault S, Dufresne L, Kurosawa R, et al. Genomic and transcriptomic analyses of aortic stenosis enhance therapeutic target discovery and disease prediction. Nat Genet. 2026 Jan;58(1):57–66. doi:10.1038/s41588-025-02417-6

30. Mishra A, Malik R, Hachiya T, Jürgenson T, Namba S, Posner DC, et al. Stroke genetics informs drug discovery and risk prediction across ancestries. Nature. 2022 Nov;611(7934):115–23. doi:10.1038/s41586-022-05165-3 PubMed PMID: 36180795; PubMed Central PMCID: PMC9524349.

31. Henry A, Mo X, Finan C, Chaffin MD, Speed D, Issa H, et al. Genome-wide association study meta-analysis provides insights into the etiology of heart failure and its subtypes. Nat Genet. 2025 Apr;57(4):815–28. doi:10.1038/s41588-024-02064-3

32. Roselli C, Surakka I, Olesen MS, Sveinbjornsson G, Marston NA, Choi SH, et al. Meta-analysis of genome-wide associations and polygenic risk prediction for atrial fibrillation in more than 180,000 cases. Nat Genet. 2025 Mar;57(3):539–47. doi:10.1038/s41588-024-02072-3 PubMed PMID: 40050429; PubMed Central PMCID: PMC12094172.

33. Sun BB, Chiou J, Traylor M, Benner C, Hsu YH, Richardson TG, et al. Plasma proteomic associations with genetics and health in the UK Biobank. Nature. 2023 Oct 12;622(7982):329–38. doi:10.1038/s41586-023-06592-6

34. Burgess S, Davey Smith G, Davies NM, Dudbridge F, Gill D, Glymour MM, et al. Guidelines for performing Mendelian randomization investigations: update for summer 2023. Wellcome Open Res. 2019;4:186. doi:10.12688/wellcomeopenres.15555.3 PubMed PMID: 32760811; PubMed Central PMCID: PMC7384151.

35. Korthauer K, Kimes PK, Duvallet C, Reyes A, Subramanian A, Teng M, et al. A practical guide to methods controlling false discoveries in computational biology. Genome Biol. 2019 Jun 4;20(1):118. doi:10.1186/s13059-019-1716-1 PubMed PMID: 31164141; PubMed Central PMCID: PMC6547503.

36. Giambartolomei C, Vukcevic D, Schadt EE, Franke L, Hingorani AD, Wallace C, et al. Bayesian Test for Colocalisation between Pairs of Genetic Association Studies Using Summary Statistics. Williams SM, editor. PLoS Genet. 2014 May 15;10(5):e1004383. doi:10.1371/journal.pgen.1004383

37. Wu T, Hu E, Xu S, Chen M, Guo P, Dai Z, et al. clusterProfiler 4.0: A universal enrichment tool for interpreting omics data. The Innovation. 2021 Aug;2(3):100141. doi:10.1016/j.xinn.2021.100141

38. Silvestri L, Pagani A, Nai A, De Domenico I, Kaplan J, Camaschella C. The serine protease matriptase-2 (TMPRSS6) inhibits hepcidin activation by cleaving membrane hemojuvelin. Cell Metab. 2008 Dec;8(6):502–11. doi:10.1016/j.cmet.2008.09.012 PubMed PMID: 18976966; PubMed Central PMCID: PMC2648389.

39. Sirlin CB, Reeder SB. Magnetic resonance imaging quantification of liver iron. Magn Reson Imaging Clin N Am. 2010 Aug;18(3):359–81, ix. doi:10.1016/j.mric.2010.08.014 PubMed PMID: 21094445; PubMed Central PMCID: PMC3430384.

40. Valenti L, Rametta R, Dongiovanni P, Motta BM, Canavesi E, Pelusi S, et al. The A736V TMPRSS6 polymorphism influences hepatic iron overload in nonalcoholic fatty liver disease. PLoS One. 2012;7(11):e48804. doi:10.1371/journal.pone.0048804 PubMed PMID: 23144979; PubMed Central PMCID: PMC3489825.

41. Handa P, Vemulakonda AL, Maliken BD, Morgan-Stevenson V, Nelson JE, Dhillon BK, et al. Differences in hepatic expression of iron, inflammation and stress-related genes in patients with nonalcoholic steatohepatitis. Ann Hepatol. 2017 Feb;16(1):77–85. doi:10.5604/16652681.1226818 PubMed PMID: 28051796.

42. Suárez-Ortegón MF, McLachlan S, Price AH, Fernández-Balsells M, Franch-Nadal J, Mata-Cases M, et al. Decreased iron stores are associated with cardiovascular disease in patients with type 2 diabetes both cross-sectionally and longitudinally. Atherosclerosis. 2018 May;272:193–9. doi:10.1016/j.atherosclerosis.2018.03.028

43. Barad A, Clark AG, Pressman EK, O’Brien KO. Associations Between Genetically Predicted Iron Status and Cardiovascular Disease Risk: A Mendelian Randomization Study. JAHA. 2024 Jun 4;13(11):e034991. doi:10.1161/JAHA.124.034991

44. Liu Y, Clarke R, Bennett DA, Zong G, Gan W. Iron Status and Risk of Heart Disease, Stroke, and Diabetes: A Mendelian Randomization Study in European Adults. JAHA. 2024 Mar 19;13(6):e031732. doi:10.1161/JAHA.123.031732

45. Quezada-Pinedo HG, Cajachagua-Torres KN, Ahanchi NS, Khatami F, Muka T, Huicho L, et al. Genetically predicted iron status and cardiovascular function and structure: a Mendelian randomization study. European Journal of Preventive Cardiology. 2026 Feb 18;33(3):394–403. doi:10.1093/eurjpc/zwaf498

46. Barrows IR, Devalaraja M, Kakkar R, Chen J, Gupta J, Rosas SE, et al. Race, Interleukin-6, TMPRSS6 Genotype, and Cardiovascular Disease in Patients With Chronic Kidney Disease. J Am Heart Assoc. 2022 Sep 20;11(18):e025627. doi:10.1161/JAHA.122.025627 PubMed PMID: 36102277; PubMed Central PMCID: PMC9683639.

47. Afsar RE, Kanbay M, Ibis A, Afsar B. In-depth review: is hepcidin a marker for the heart and the kidney? Mol Cell Biochem. 2021 Sep;476(9):3365–81. doi:10.1007/s11010-021-04168-4 PubMed PMID: 33942218.

