## Supplemental Materials for "Human genetic investigation of TMPRSS6 as a therapeutic target for liver disease: efficacy and safety liabilities"

**Supplemental Figure 1**

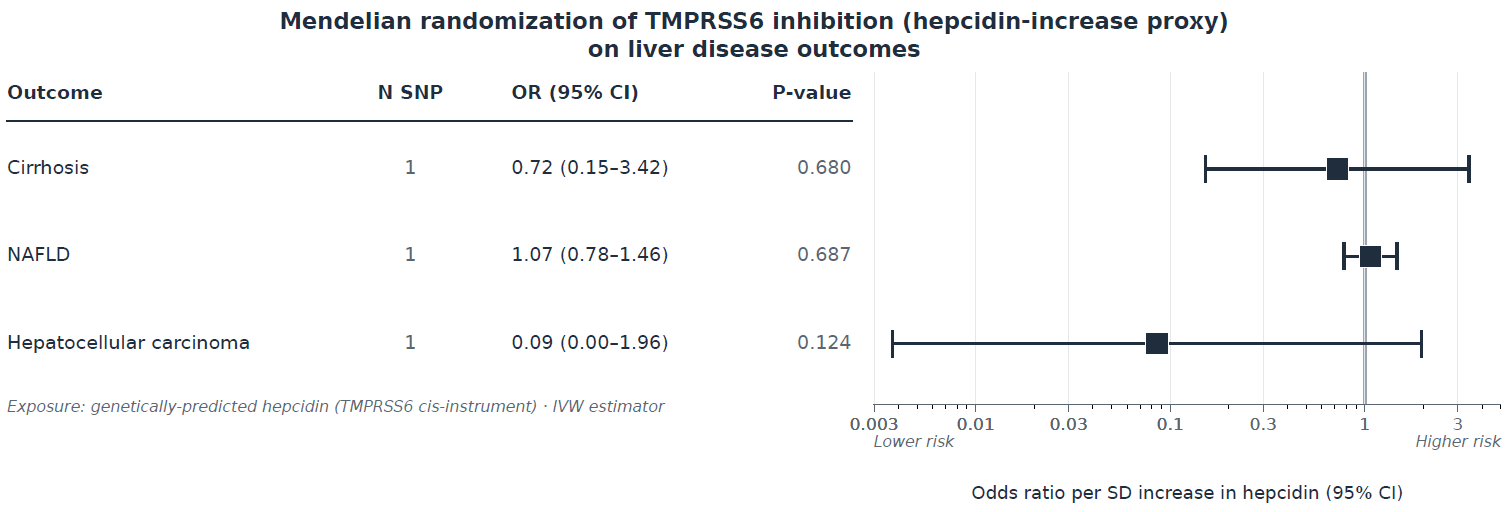

**Supplemental Figure 2**

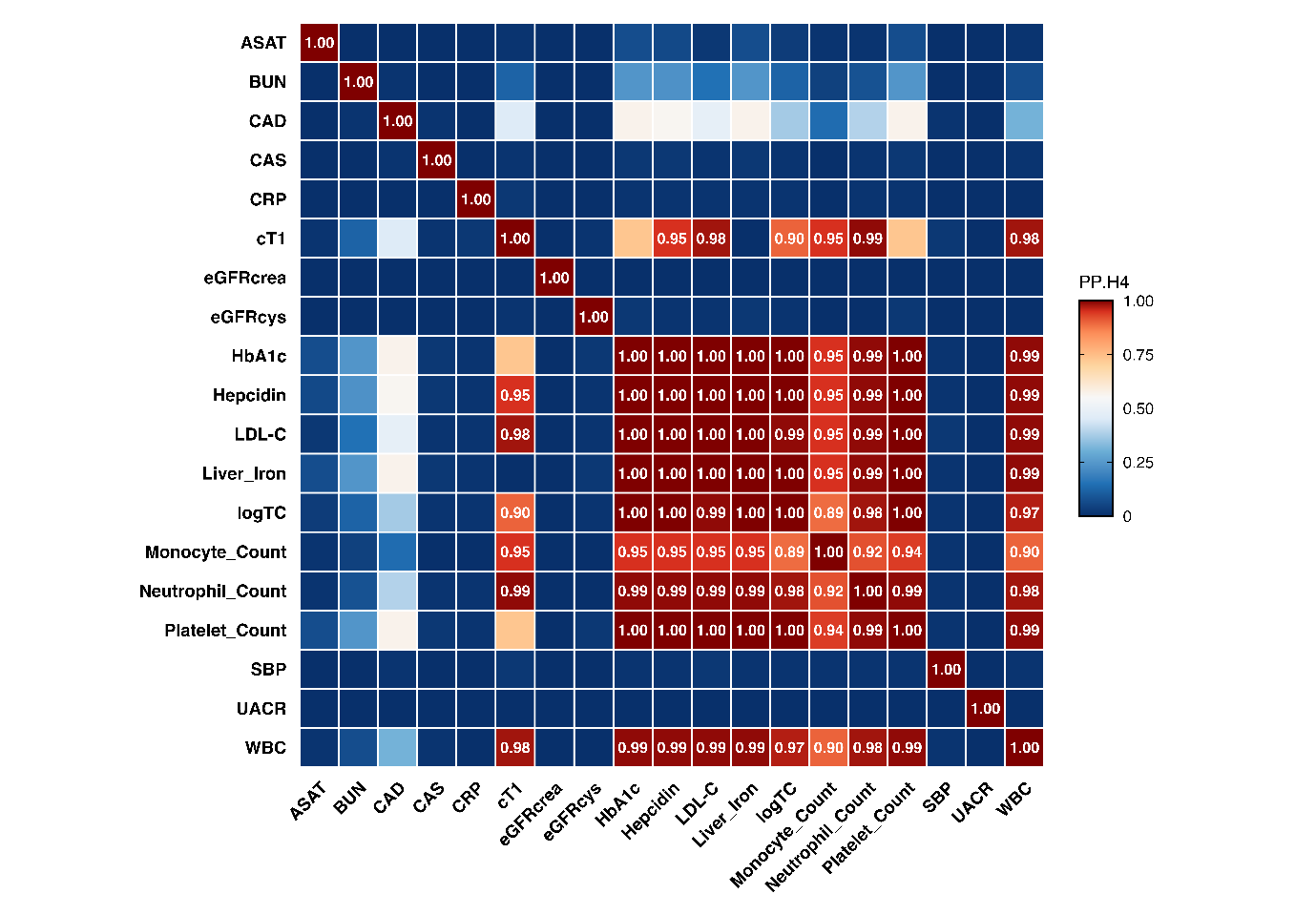

**Supplemental Figure 2**

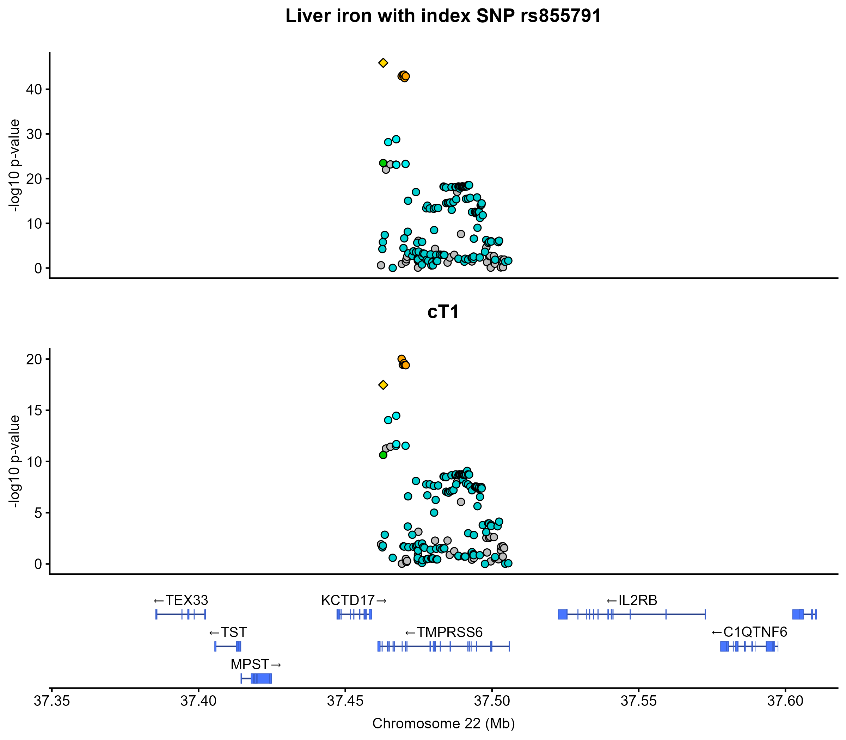
